# Genetic Prediction of Parkinson’s Diagnosis: Firth to Ensemble Learning

**DOI:** 10.64898/2026.07.11.26357847

**Authors:** Michael J. Espero

**Author notes:** Correspondence concerning this should be addressed to Michael J. Espero.

## Abstract

By utilizing a targeted genetic assay within a Fox Insight cohort (*N* = 1,987), this research establishes a hybrid, transparent, and interpretable predictive framework. Initial modeling via Firth penalized logistic regression discovered enrichment regarding the GBA N370S locus, highlighting the critical role of epidemiological evaluation in human study populations. Advanced ensemble learning methods, refined through a meta-learner gradient boosting machine, attained an *out-of-sample* AUC of 0.929 and MCC of 0.743 on 15% of the analysis dataset partitioned via random sampling and strictly held-out from model training. Both global, visual machine learning explanations and local-Shapley interpretations provide *transparency* into the models and individual predictions representative of practical, collaborative human-artificial intelligence efforts, offering a solution that supports classification while remaining accessible and economical.

## Introduction

Since the landmark work of Chang et al. (2017) and Nalls et al. (2019), even more strides have been made to identify heritable risk for Parkinson’s disease (PD), the world’s second most prevalent neurodegenerative disease. The former found 17 novel risk loci for the heritability of PD in a sample of 26,035 cases and 403,190 controls. The latter work identified 38 novel risk signals in 37 loci in a sample of 37,700 cases; 18,600 first-degree relatives (proxy-cases), and 1,400,000 controls. Tolosa, Garrido, Scholz, and Powe (2021) suggest over 90 risk loci, including common and rare variants that funnel into causal biological pathways: interrupted homeostasis of mitochondria, cell necrobiology, inflammatory signaling, and intracellular transport. Many of the *genetic* sources of risk identified are found to be associated with brain-first (rather than body-first) decline, making them among the most reasonable features to investigate when attempting to detect neurocognitive risk, which may be less directly visible to the clinician (Passaretti et al., 2025). While suggesting around 10 genetic precipitates to Parkinsonian pathology, Bloem, Okun, and Klein (2021) assert that relevant heterogeneity justifies a clinical-research imagination for over 6 million variations of Parkinson’s disease throughout the world.

With machine learning (ML) methods and Fox Insights data, Rahman and Liu (2023) developed models classifying those with PD from those without. The ML models performed up to an AUC of 0.74. Given precedence for genetic, environmental (i.e. pesticide and herbicide exposure), and age-related risk factors (Tenchov, Sasso, and Zhou, 2025), multifaceted approaches are called for to assist clinical assessments and provide large-scale support for the early identification of risk- economically and whenever possible. The present work leverages classical statistical and ML models to predict PD diagnosis/risk (Gottesman et al., 2024) and explain sources of risk, including genetic associations of PD risk based on a targeted genetic assay.

### On Diagnosis

The processes of identifying phenomena thought to precipitate disease bring with them a host of assumptions and conceptual constructs. To move forward without consideration of the tenability of these foundations and into quantitative classification or clinical reflection undermines not only the epistemological sovereignty of the people observed, but overlooks grounds that may contain some clues we need to reconsider as we develop our understanding. Falcato (2025) reflects on Kant’s work including the ‘*Essay on the Maladies of the Head’*, where the situated nature of maladies affecting psychology are considered. In a view so expressed, main *genera* may be differentiated by (at least) as many mental faculties as are observed/experienced to be afflicted. Thus, one might suggest that an approach to classifying disease may be reliant upon characterizing the specific and co-compromised physical systems and doubly so, the psychological capacities that emerge (or are hindered so) from them. Beyond Kant’s notions of imagination, understanding, and reason - capacities for a steady stance & gait (for instance), broad cognitive abilities, their differentiated and oblique functional elements, as well as experienced psychological qualities of daily life - may be formative of the stages of many attractor states thought to encompass disease.

Scadding (1988) argues that as knowledge advances, syndromal characterization (clinical description as the ‘starting point’ with reason for dissension) of disease is displaced by aetiology (when possible). Rather than medical and research practice progressing by the basis of diagnosis and treatment specified by textbooks or other authority, it is the *discovery* and evermore elucidated and *revised* nature of causal factors (and networks) and recognition of functional and/or structural disorders that provide weight to our contributions and the knowledge required for holistic treatment (explanation, support, relief, and amelioration) of those who suffer. Scadding (1988) adds that the positioning of a group relative to a population in terms of biological disadvantage behooves disease classification.

Friedman (2024) develops philosophical scaffolding unyoked from a specific discipline, providing a pluralistic framework in which to analyze medical concepts such as the notion of malady. By imagining a binocular framework, Friedman suggests the simultaneous vision of notions regarding health and malady including a lens of aspects (bioscientific, individual, and collective) and a lens of perspectives (biomedical, first-person, and social). From this view, a comprehensive and non-hierarchical approach to understanding complexity relevant to health, malady, and human experience emerges, offering epistemic pluralism without commitment to any single disciplinary element. Powell and Scarffe (2019) concur, insofar as notions of disease must involve both biological and moral objectivity if they are to survive their situatedness in, for instance, healthcare institutions. From these philosophical approaches to the definition of disease, we turn to the diagnosis of Parkinson’s disease, specifically, before moving on to an empirical classification (and evaluation thereof) - aiming to characterize the global (given the participant group) phenotype underlying the pool’s clinical distinction as well as explanatory variation present among those people assumed classifiable as cases or controls (local machine learning interpretability).

### Parkinson’s

In reality, human experiences of suffering and distinction of disease from healthy states exist in much greater variation than binary classification (or even stages), taken at face value, might suggest. The cardinal clinical signs of Parkinson’s disease, among a spectrum of motor and non-motor features - are thought to include tremor (even at rest), bradykinesia (compromised speed of movement), loss of postural reflexes, neurocognitive dysfunction, and pain (Jankovic, 2007). Additionally, a strong indication of Parkinson’s disease is thought to involve response to levodopa (improvement with treatment considered *suggestive* of PD), given repeated observations of neuronal atrophy localized to the substantia nigra pars compacta (Bloem, Okun, and Klein, 2021).

Non-motor symptoms of PD are relatively more difficult to recognize and ensure adequate treatment (Chaudhuri, Healy, and Schapira, 2006; Bloem, Okun, and Klein, 2021). Simultaneously, non-motor symptoms *may* be the most burdensome and negatively impactful in quality of life, including (but not limited to) sleep disturbance, difficulties in facets of memory, as well as what may be experienced as episodes of confusion, depression, and anxiety (Tenchov, Sasso, and Zhou, 2025). The severity of sleep pathologies, for instance, ranges from everyday compromised health (psychophysiological) to, at the acute-extreme, traffic accidents caused by sudden sleep onset.

With regard to Parkinson’s disease and other neurodegenerative conditions, it must not be overlooked that individuals who face varied constellations of biological phenomena including (but not limited to) dopaminergic neuronal dysfunction and pathological alpha synuclein species (Höglinger & Lang, 2024) also have varied *experiences* (see Bennett & Hacker, 2022) that impact their day-to-day lives and well-being. Chadhuri et al. (2006) and Simuni (2025) assert that despite great progress in diverse assessment tools drawn by even the largest of relevant research organizations, researchers and clinicians are in need of new scales and approaches for understanding and detecting multifaceted conditions (such as Parkinson’s), asserting that openness to heterogeneity of hypothesized biological stages as well as the emergence of new biomarkers should temper determinations of data-driven definitions. While disease is a complex spectrum, binary classification via statistical and machine learning is a necessary pragmatic step for triage and risk prioritization. The present work provides economical, deployable, transparent classifiers to support clinical practice (*the classifier framework may be generalizable to other groups and settings*, however, the results are appropriate to refer to the specific Fox Insight sample).

## Methods

### Participants

Data was sourced from Fox Insight participant groups. The analysis sample (Appendix A) included 1,987 Fox Insight participants (855 women and 1,132 men) with 1,861 people diagnosed with PD (cases) and 126 controls (no PD diagnosis). Fox Insights participants averaged 64 years of age, agreeing to volunteer genetic, questionnaire, and other health data. The Fox Insight (data) dictionary was used to ensure valid encoding of analysis data.

### Genetic Reference

To address appropriate baseline state assignment for statistical inference, wild-type alleles for the genetic variants were validated using the NCBI Database of Short Genetic Variations. Ancestral alleles on the forward strand (GRCh38 assembly) served as the non-carrier reference. Within this framework, reference homozygote genotypes were encoded as *TT* for GBA N370S (rs76763715; i4000415), *GG* for LRRK2 G2019S (rs34637584), *CC* for GBA E326K (rs2230288), *CC* for GBA T369M (rs80356773/rs75822236), and *GG* for PRKN R275W (rs34424986). These validated encodings were initially applied as baselines (explicitly, ensuring calculated odds ratios accurately reflect the influence of risk variant carrier status (Sherry et al., 2001), however, the majority variants *observed in the sample* were operationalized as reference encodings, anchoring the regression against the most frequent homozygote.

### Statistical Analysis and Machine Learning

All statistical analyses and data preprocessing were conducted in R. The primary outcome was a binary classification of Parkinson’s disease diagnosis (0 as Control, 1 as Case). Covariates included in the adjusted models were age, biological sex (with XX/Female designated as the reference baseline), and handedness.

Outcome:

> *“Do you currently have a diagnosis of Parkinson’s disease, or parkinsonism, by a physician or other health care professional?”*

**A multivariate Firth penalized logistic regression model** was fit to estimate the adjusted odds ratios (*OR*) and 95% confidence intervals (*CI*) for Parkinson’s disease diagnosis. The Firth penalized likelihood approach was chosen with the aim of minimizing the risk of biased parameter estimates arising from the sparse data structures typical of rare genetic variants and also to compare with machine learning/artificial intelligence methods. To account for multiple comparisons across the genetic terms, *p*-values were adjusted using the Benjamini-Hochberg false discovery rate (FDR) procedure. Finally, the multivariate model’s ability to predict the clinical Parkinson’s diagnoses in the analysis sample was assessed using the area under the receiver operating characteristic curve (AUC), the Brier score, and logarithmic loss.

Rare variants were explicitly coded non-carrier vs. carrier (‘Non-carriers’ assumed as i4000415: TT; rs34637584: GG; rs2230288: CC; rs80356773: CC; rs34424986: GG). Odds ratios (OR), Wald tests with confidence intervals, frequentist p-values, and false-discovery rates (FDR) are provided.

Transparency into **machine learning models** is achieved by the inclusion of global variable importance permutation plots (*Which variables influence how the model understands the data, on the whole, best?*), individual conditional expectations plots (*How does the range of a feature influence predictions, holding all other variables constant?*), and local-Shapley Fox Insights participant (de-identified) prediction-level (individual-level) plots (*How do we explain why a prediction was made for an individual?*). Additionally, *predicted* PD (diagnosis) probability is provided given the suites of machine learning models for individual case (PD) and control (no PD diagnosis) participants; this is intended to convey the relative differences in per-model-confidence in predicting the apparent clinical disease classification and for this work to demonstrate predictive heterogeneity when considering global model performance and individual-level prediction.

The analysis data was separated randomly by a 70/15/15 split to allocate 70% of the data to training, 15% to validation, and 15% as an unseen, separate testing sample. After the model-building process using the training and validation datasets the separate testing dataset was used with the model to assess generalizability to unseen data. An ensemble was fit in 6 minutes in Java, leveraging a remote NVIDIA T4 GPU on the analysis data. During the build process, six models were chosen for the ensemble including a gradient boosting machine (GBM), an extreme gradient boosting machine (XGB), a distributed random forest (DRF), a deep neural network (DNN), and a generalized linear model (GLM). Following the fit of these base learners, a meta-learner gradient boosting machine was applied, fitted on all base learner predictions (not the input features). The build process was performed a second time for 6 minutes. In the second build process, a single GBM outperformed all ensembles.

Additionally, a generalized low rank model (GLRM) was fit on the training data set, *A*, (80%) with rank 4. Demography space (the X-matrix) was extracted and added to the training dataset to augment input features in aim of improved predictive ability (the width of *A* widened by rank 4 denoised, continuous columns). A series of machine learning models were fitted, leveraging the *denoised* columns derived from the GLRM in addition to the original input features. The GLRM was applied to extract and concatenate the X-matrix features as augmentation to the validation and testing sets (separately), as well.

## Results

### Statistical Model Performance

Overall, the multivariate model demonstrated strong predictive performance (Table 1, below) on the analysis sample, yielding an area under the receiver operating characteristic curve (AUC) of 0.905 and a Brier score of 0.028. Full parameter estimates, including unadjusted *p*-values and false discovery rate (FDR) adjusted *q*-values for all genetic terms, are detailed in Table 2. Visual representation of the log-transformed odds ratios and corresponding confidence intervals is provided in Figure 1.

**Table 1.**

| Model Performance |  |  |  |
| --- | --- | --- | --- |
| Firth Logistic Regression on the Analysis Sample |  |  |  |
| Log Loss | Brier | AUC | N |
| 0.118 | 0.028 | 0.905 | 1987 |

### Statistical Analysis

**Figure 1:**
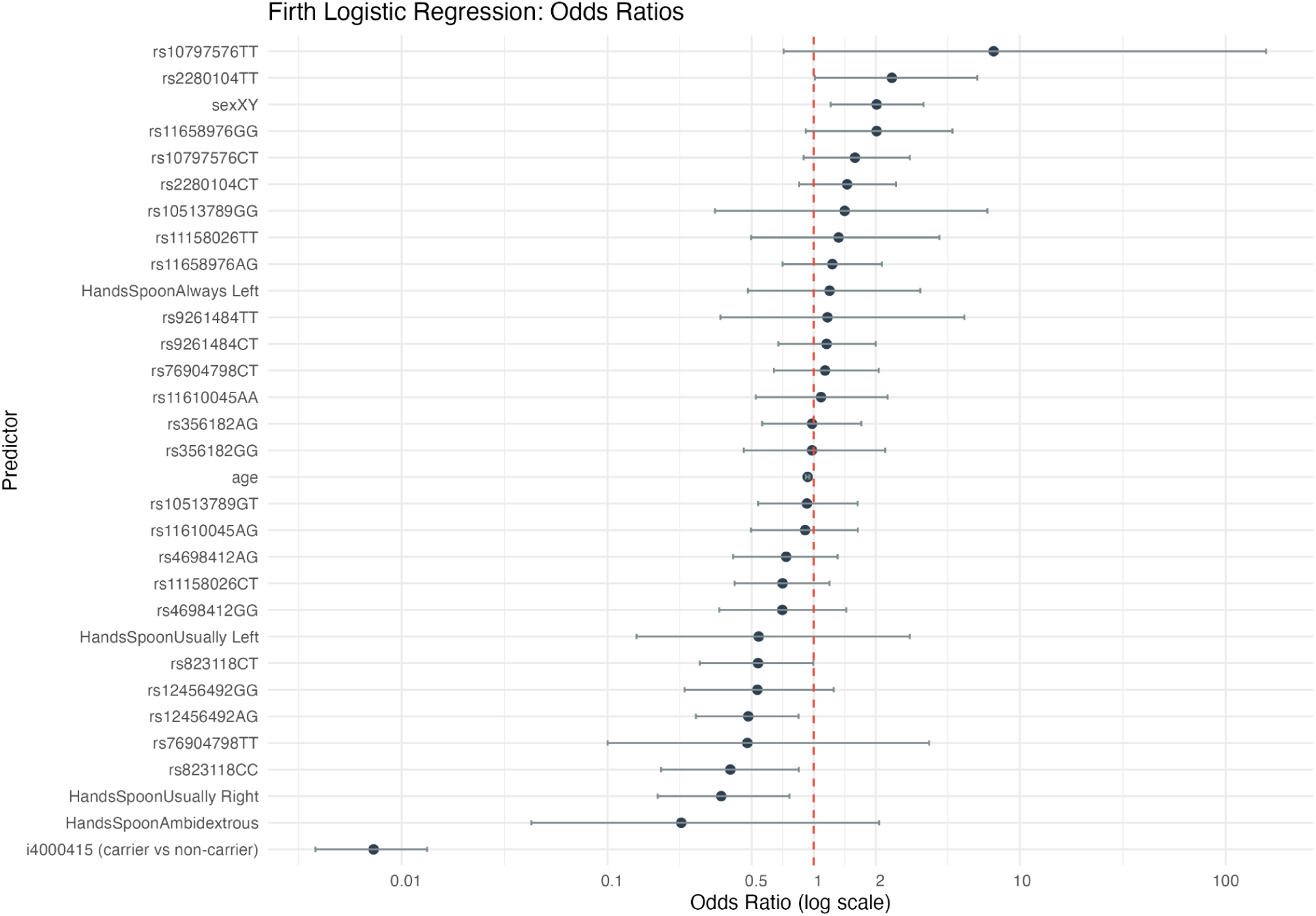

A multivariate Firth penalized logistic regression model provides estimates of adjusted odds ratios (*OR*) for Parkinson’s disease diagnosis *in-sample*, adjusting for covariates of age (centered), biological sex (females, *XX*, as the reference), and handedness (‘always right’-handed spoon usage as the reference). As expected for this disease pathology, demographic covariates were associated with diagnosis; male sex (*XY*) was significantly associated with increased odds of diagnosis (*OR* = 2.02, 95% CI [1.21, 3.41], *p* = .007), indicating that males have approximately twice the odds of a PD diagnosis compared to the female baseline in the analysis sample. The Firth penalized likelihood method was selected *a priori* to mitigate estimation bias inherent in sparse data structures and to resolve issues of complete separation typical of rare genetic variants^1^.

The Firth approach enabled sample modeling across all included variants (see Table 2) with adequate cell counts. Carrier status for the GBA N370S variant (i4000415) emerged with a highly significant, albeit inverted, protective association (*OR* = 0.01, 95% CI [0.00, 0.01], *p* < .001, FDR < .001). This reflects *a distinct cohort recruitment enrichment rather than what should be assumed as true biological protection in the population*. The estimate for variant rs12456492 minor allele carrier status suggests a relative protective effect, (*OR* = 0.48, 95% CI [0.27, 0.84], *p* = .010). Statistically significant protective and risk (respectively) associations were observed for rs823118 (OR = 0.39, CI [0.18, 0.85], *p* = .017) and rs2280104 (*OR* = 2.40, 95% CI [1.01, 6.23], *p* = .046). Notably, *none* of these variants withstood the Benjamini-Hochberg false discovery rate correction.

**Table 2.**

| Multivariate Firth Logistic Regression Results |  |  |  |  |  |  |  |
| --- | --- | --- | --- | --- | --- | --- | --- |
| Odds Ratios for Parkinson's Disease Diagnosis (Sample N = 1987) |  |  |  |  |  |  |  |
| Predictor | Beta | Odds Ratio | Lower 95% CI | Upper 95% CI | P-Value | Genetic Variant | FDR (q-value) |
| **i4000415 (carrier vs non-carrier)** | **-4.92** | **0.01** | **0.00** | **0.01** | **0.000** | **TRUE** | **0.000** |
| **rs12456492AG** | **-0.73** | **0.48** | **0.27** | **0.84** | **0.010** | **TRUE** | **0.131** |
| **rs823118CC** | **-0.93** | **0.39** | **0.18** | **0.85** | **0.017** | **TRUE** | **0.143** |
| **rs2280104TT** | **0.87** | **2.40** | **1.01** | **6.23** | **0.046** | **TRUE** | **0.240** |
| **rs823118CT** | **-0.62** | **0.54** | **0.28** | **0.99** | **0.048** | **TRUE** | **0.240** |
| rs11658976GG | 0.70 | 2.02 | 0.91 | 4.71 | 0.083 | TRUE | 0.346 |
| rs10797576TT | 2.01 | 7.47 | 0.71 | 156.61 | 0.106 | TRUE | 0.367 |
| rs10797576CT | 0.46 | 1.59 | 0.89 | 2.92 | 0.117 | TRUE | 0.367 |
| rs12456492GG | -0.63 | 0.53 | 0.24 | 1.25 | 0.145 | TRUE | 0.402 |
| rs2280104CT | 0.37 | 1.45 | 0.85 | 2.51 | 0.172 | TRUE | 0.430 |
| rs11158026CT | -0.35 | 0.71 | 0.41 | 1.19 | 0.194 | TRUE | 0.440 |
| rs4698412AG | -0.31 | 0.73 | 0.41 | 1.31 | 0.296 | TRUE | 0.617 |
| rs4698412GG | -0.35 | 0.70 | 0.35 | 1.44 | 0.332 | TRUE | 0.639 |
| rs76904798TT | -0.74 | 0.48 | 0.10 | 3.63 | 0.444 | TRUE | 0.768 |
| rs11658976AG | 0.21 | 1.23 | 0.71 | 2.14 | 0.461 | TRUE | 0.768 |
| rs11158026TT | 0.28 | 1.32 | 0.50 | 4.07 | 0.594 | TRUE | 0.879 |
| rs9261484CT | 0.14 | 1.15 | 0.67 | 2.00 | 0.603 | TRUE | 0.879 |
| rs10513789GG | 0.35 | 1.41 | 0.33 | 6.96 | 0.660 | TRUE | 0.879 |
| rs76904798CT | 0.13 | 1.14 | 0.64 | 2.07 | 0.668 | TRUE | 0.879 |
| rs11610045AG | -0.10 | 0.91 | 0.50 | 1.63 | 0.752 | TRUE | 0.899 |
| rs10513789GT | -0.08 | 0.93 | 0.54 | 1.63 | 0.789 | TRUE | 0.899 |
| rs9261484TT | 0.15 | 1.17 | 0.35 | 5.39 | 0.820 | TRUE | 0.899 |
| rs11610045AA | 0.08 | 1.09 | 0.52 | 2.28 | 0.827 | TRUE | 0.899 |
| rs356182AG | -0.02 | 0.98 | 0.56 | 1.70 | 0.949 | TRUE | 0.963 |
| rs356182GG | -0.02 | 0.98 | 0.46 | 2.22 | 0.963 | TRUE | 0.963 |
| **age** | **-0.07** | **0.93** | **0.92** | **0.95** | **0.000** | **FALSE** | **—** |
| **sexXY** | **0.70** | **2.02** | **1.21** | **3.41** | **0.007** | **FALSE** | **—** |
| **HandsSpoonUsually Right** | **-1.03** | **0.36** | **0.17** | **0.76** | **0.009** | **FALSE** | **—** |
| HandsSpoonAmbidextrous | -1.48 | 0.23 | 0.04 | 2.08 | 0.180 | FALSE | — |
| HandsSpoonUsually Left | -0.61 | 0.54 | 0.14 | 2.92 | 0.448 | FALSE | — |
| HandsSpoonAlways Left | 0.18 | 1.19 | 0.48 | 3.29 | 0.715 | FALSE | — |

### ML Model Performance

Global performance indices are reported based on the GBM (build group 2) performance on the unseen data (15% of the analysis sample).

| AUC | Gini | Log Loss | MCC |
| --- | --- | --- | --- |
| 0.929 | 0.859 | 0.108 | 0.743 |

Of the 290 held-out participants for the testing sample, 6 controls were misclassified (false positives) out of the 280 controls. Of the 17 cases in the testing sample, 3 were misclassified (17.6% false-negatives). By evaluating all four quadrants of the confusion matrix, the MCC attained a value of 0.743, which confirms *excellent discriminatory power* and balanced classification reliability. This suggests the framework remains robust across both the heavily represented majority class (individuals with diagnosed Parkinson’s) and the relatively sparse minority class (apparent controls).

### Visual ML Explainability

#### Variable Importance

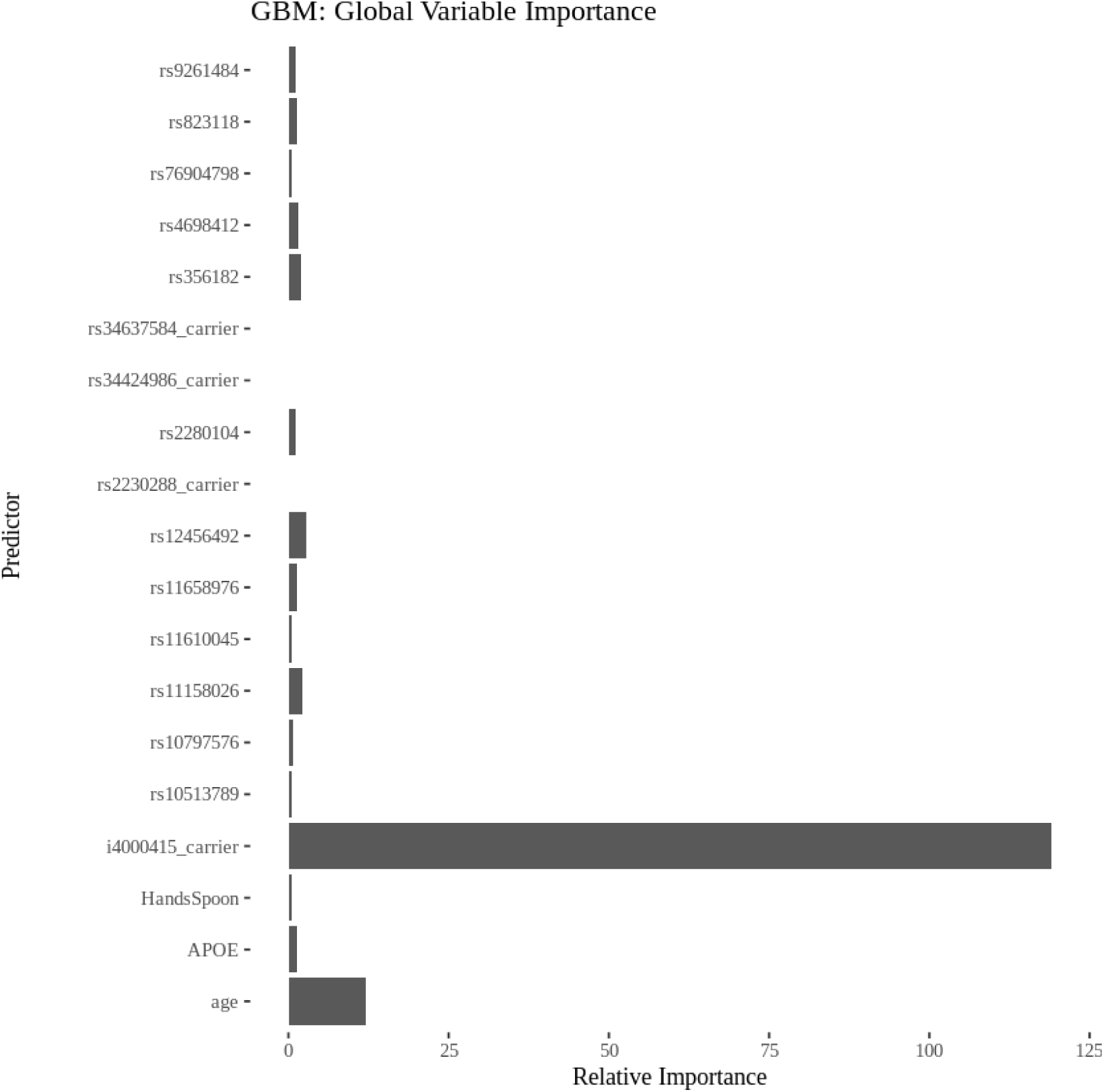

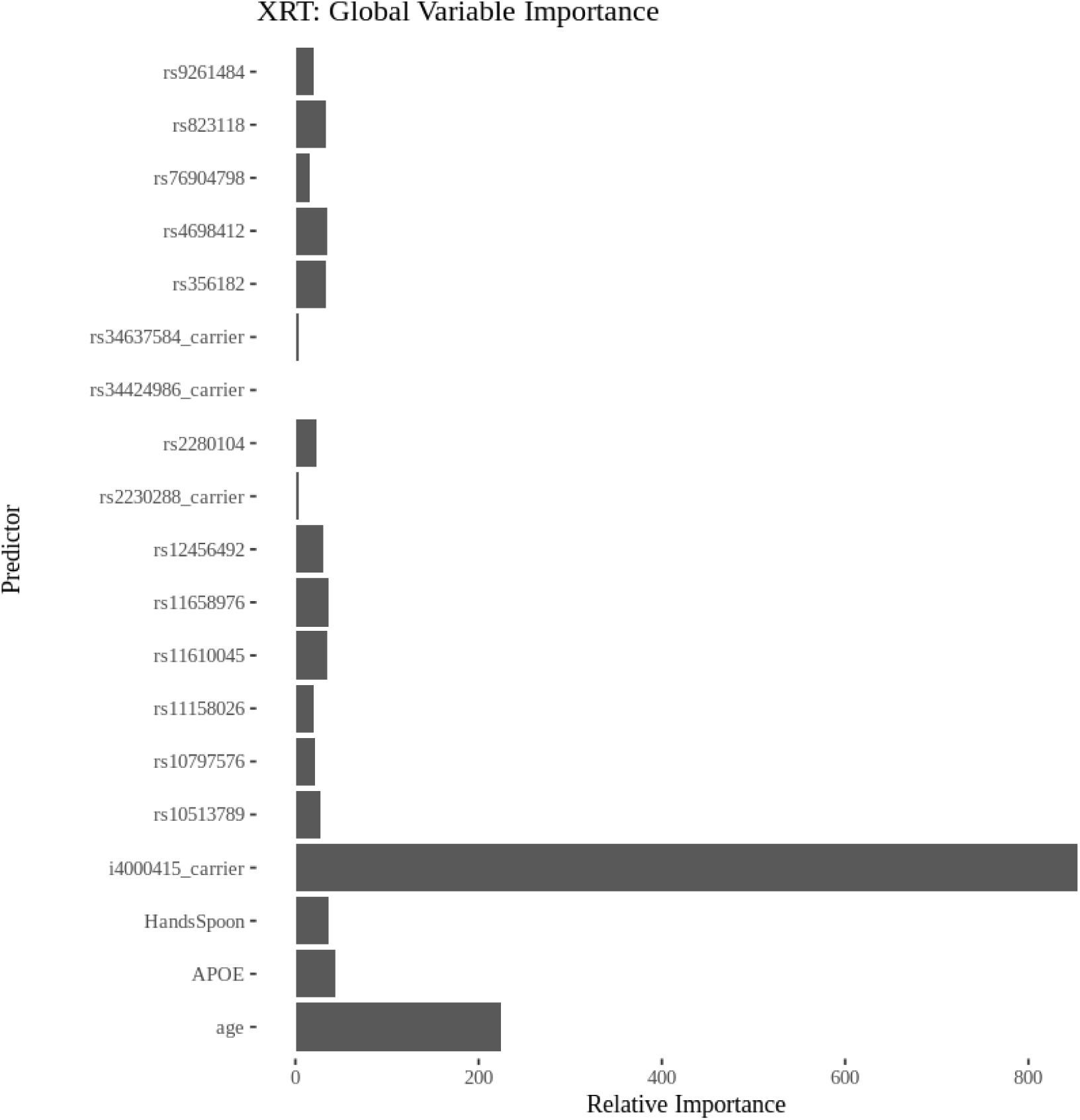

#### Individual Conditional Expectations (ICE)

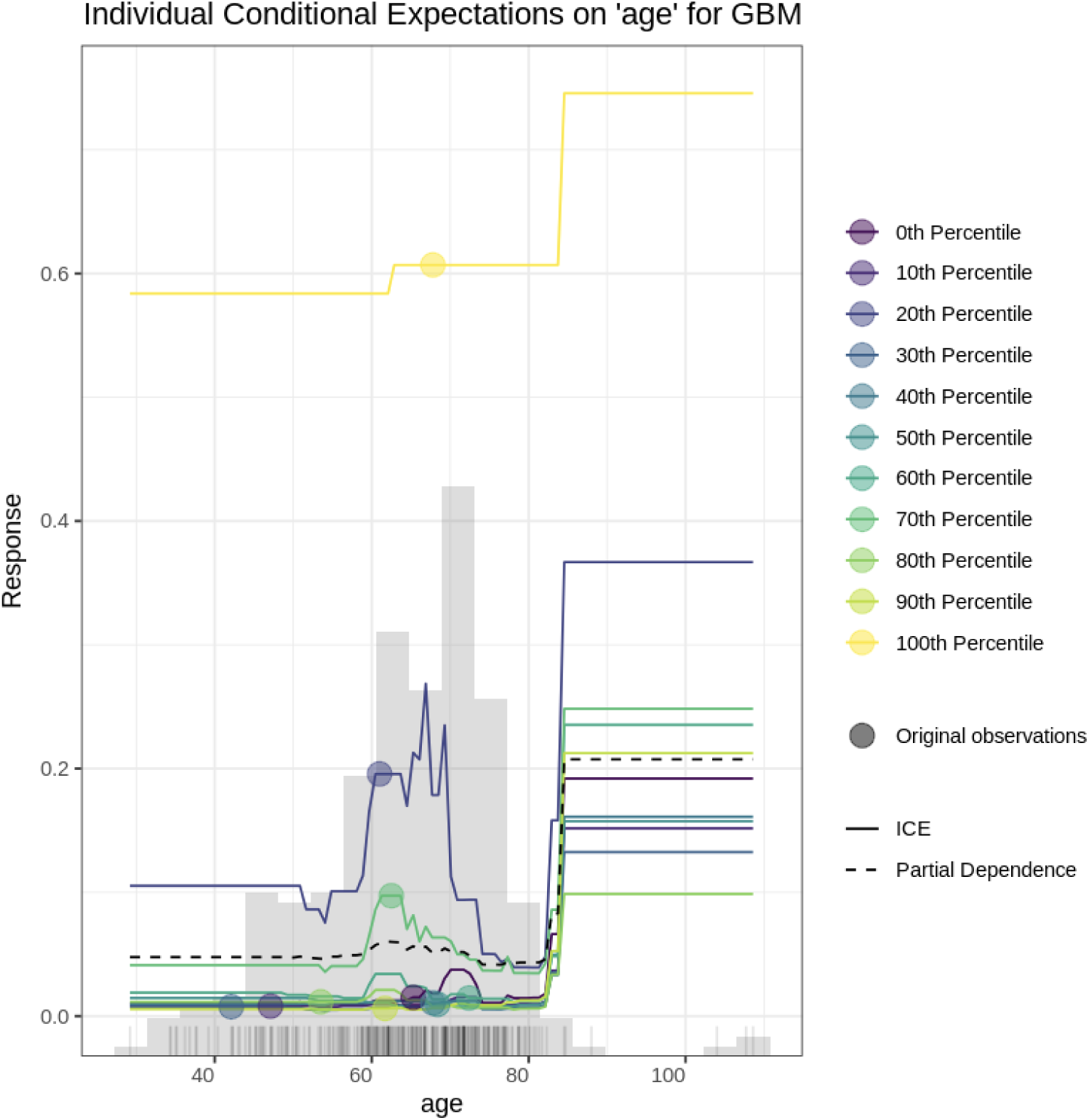

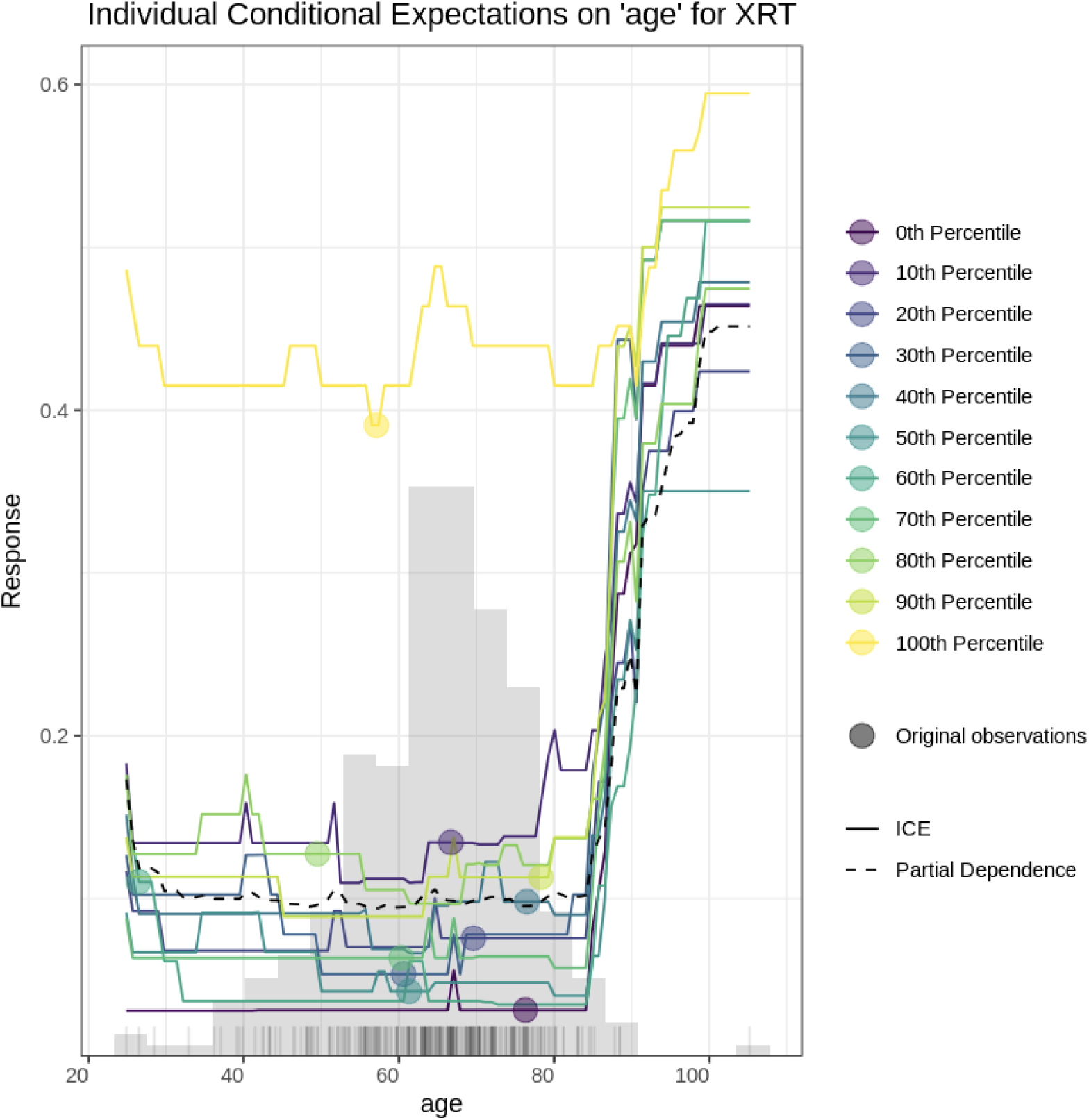

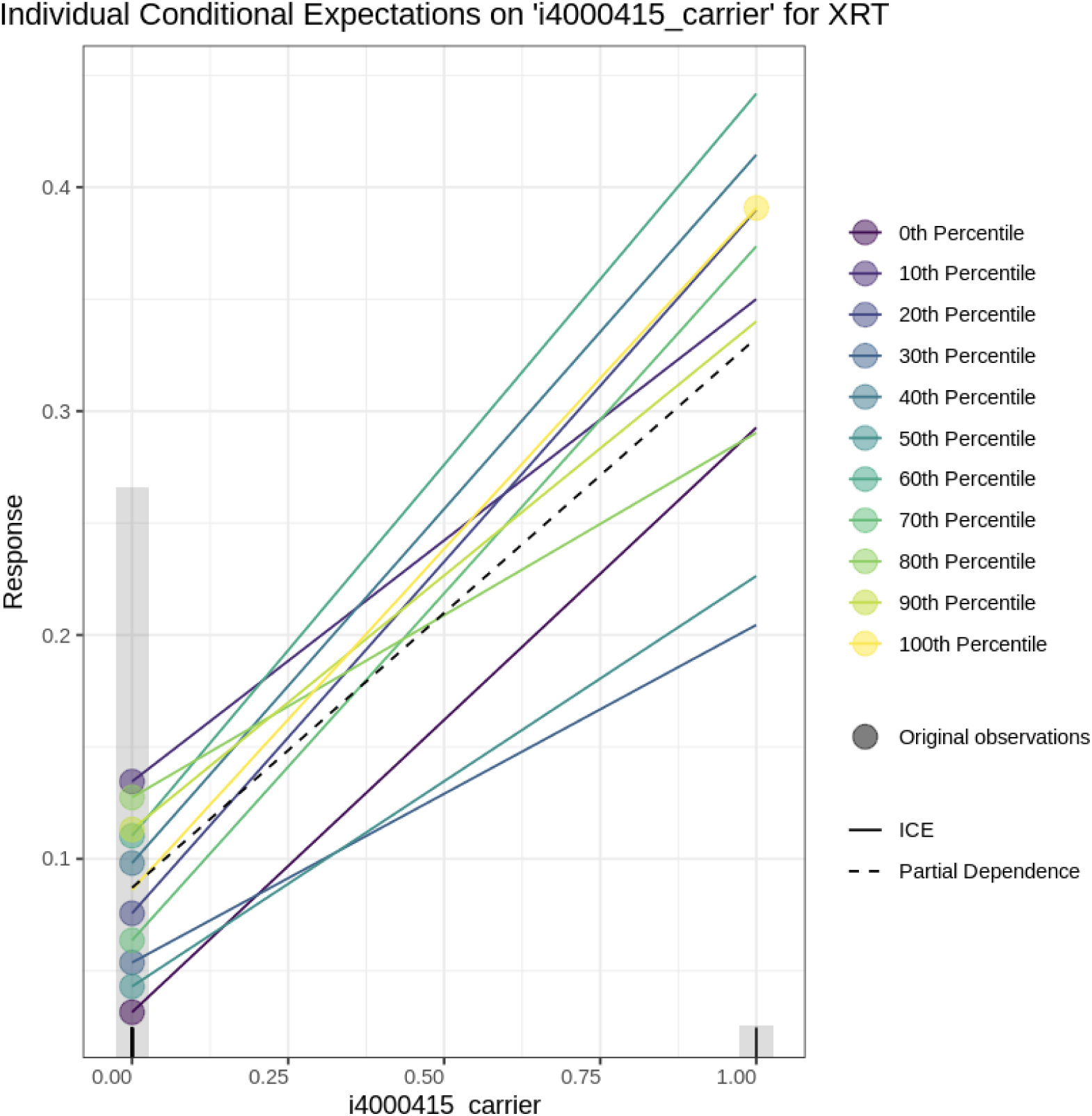

### Local Explainability

#### GBM1

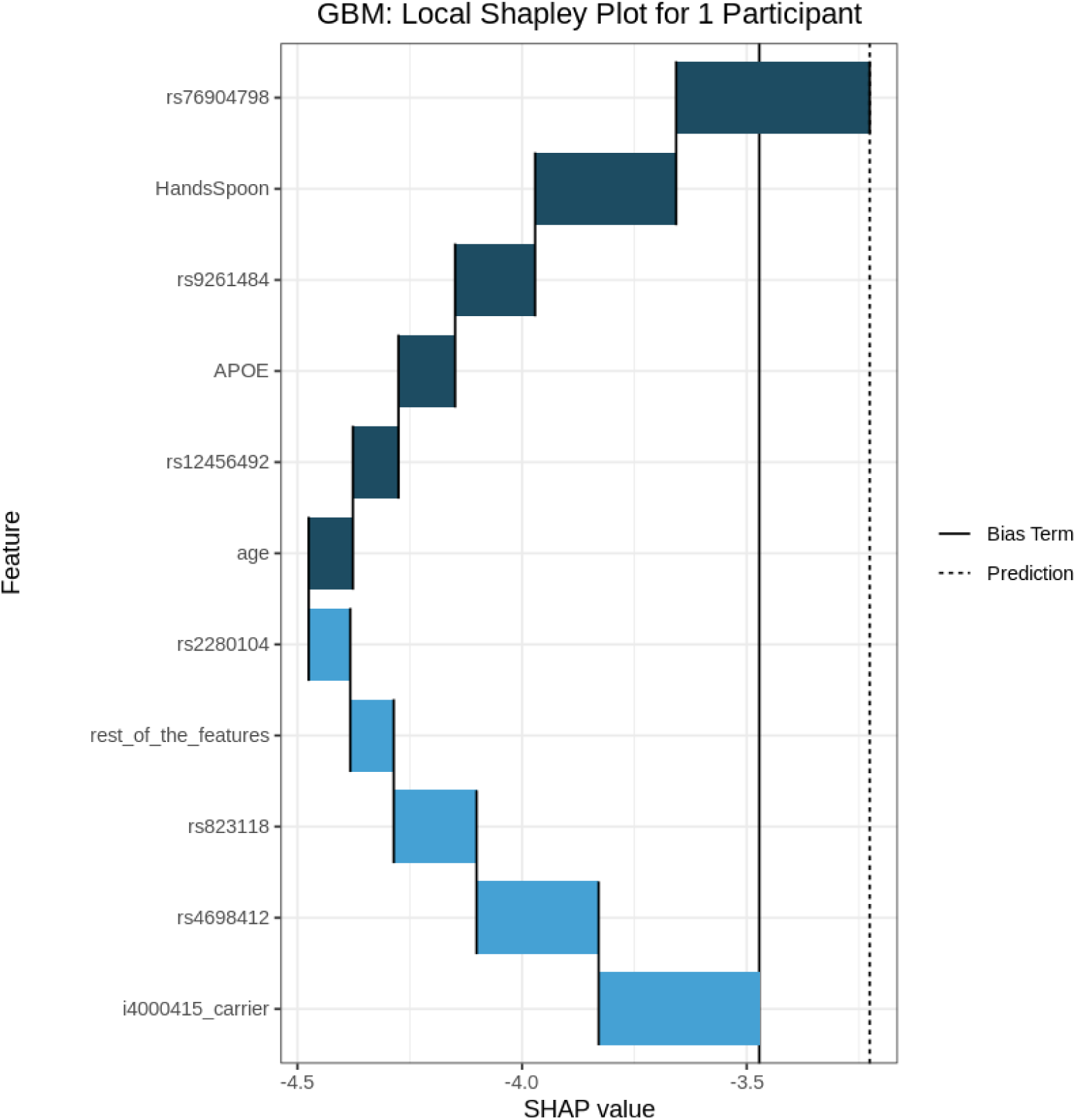

#### GLM

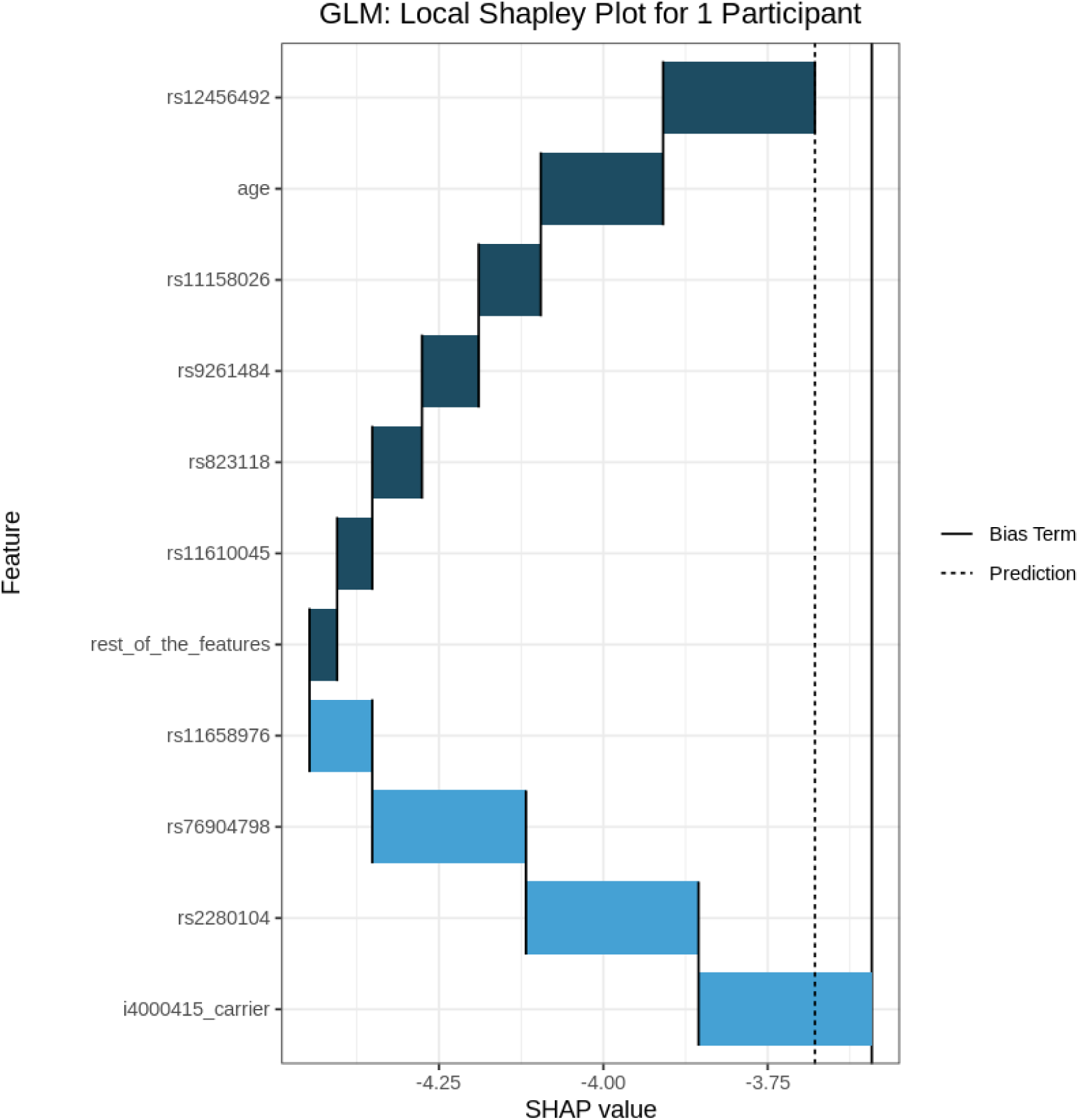

#### XRT

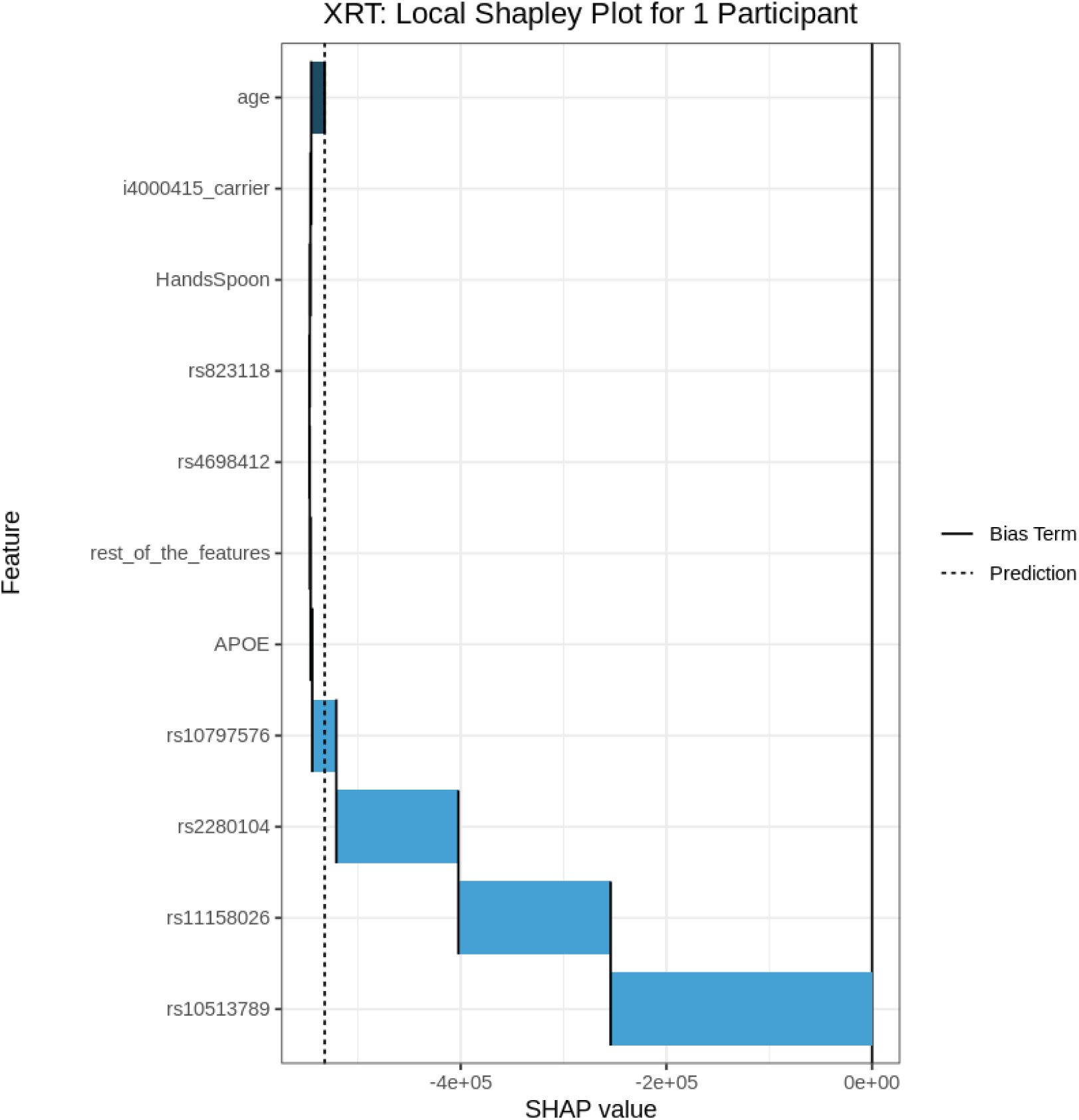

#### DRF

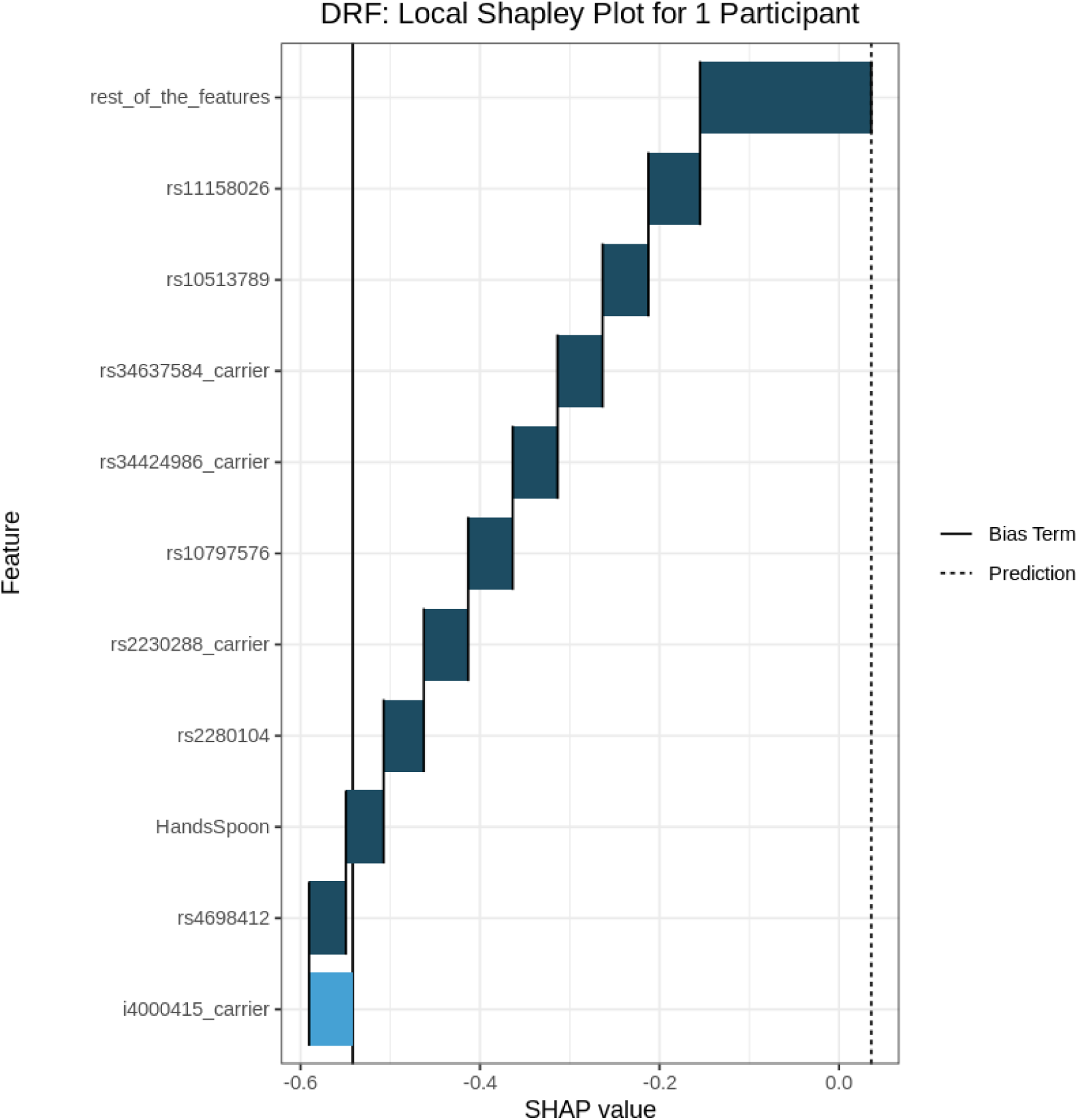

#### GBM

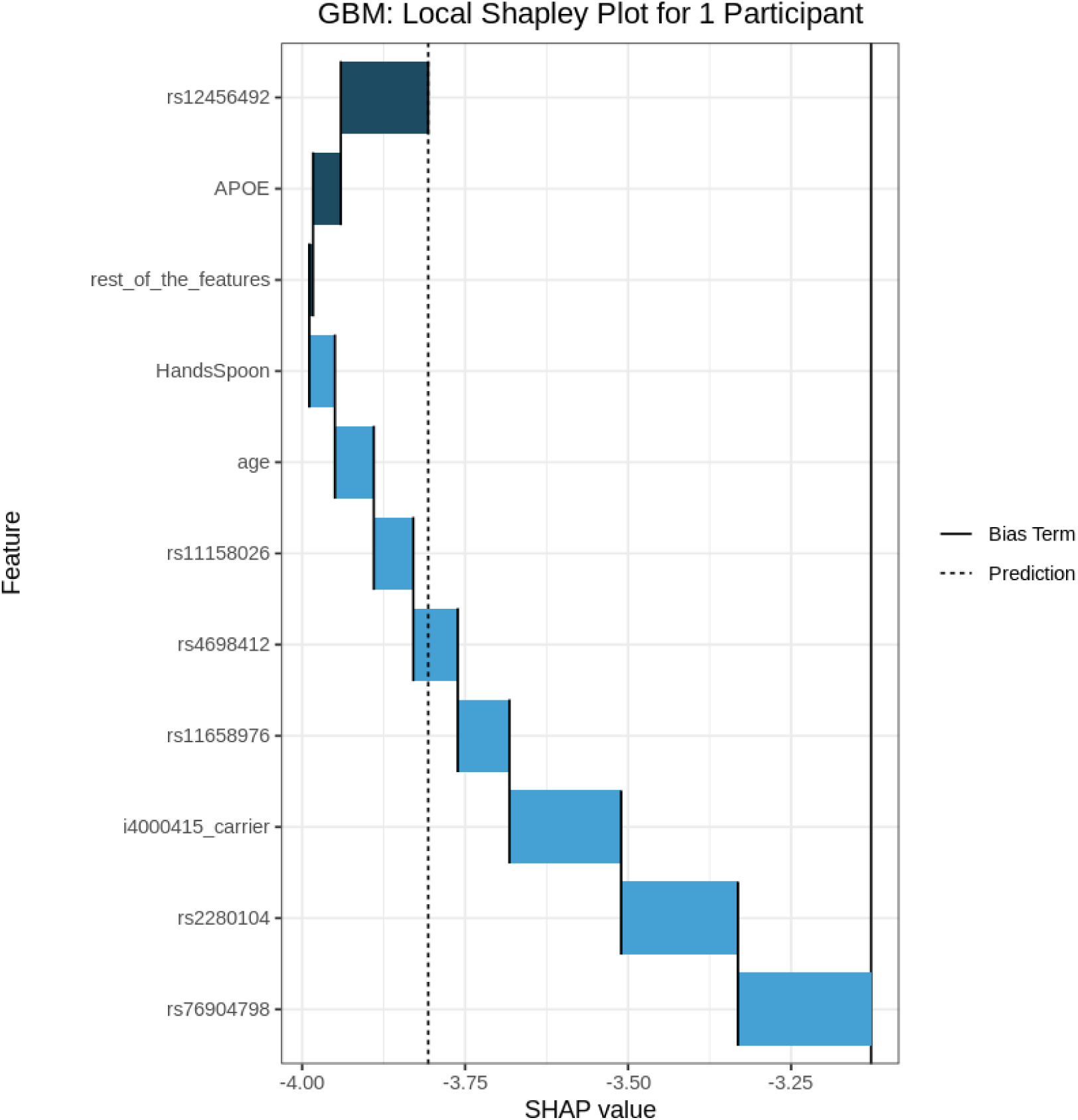

#### XGB

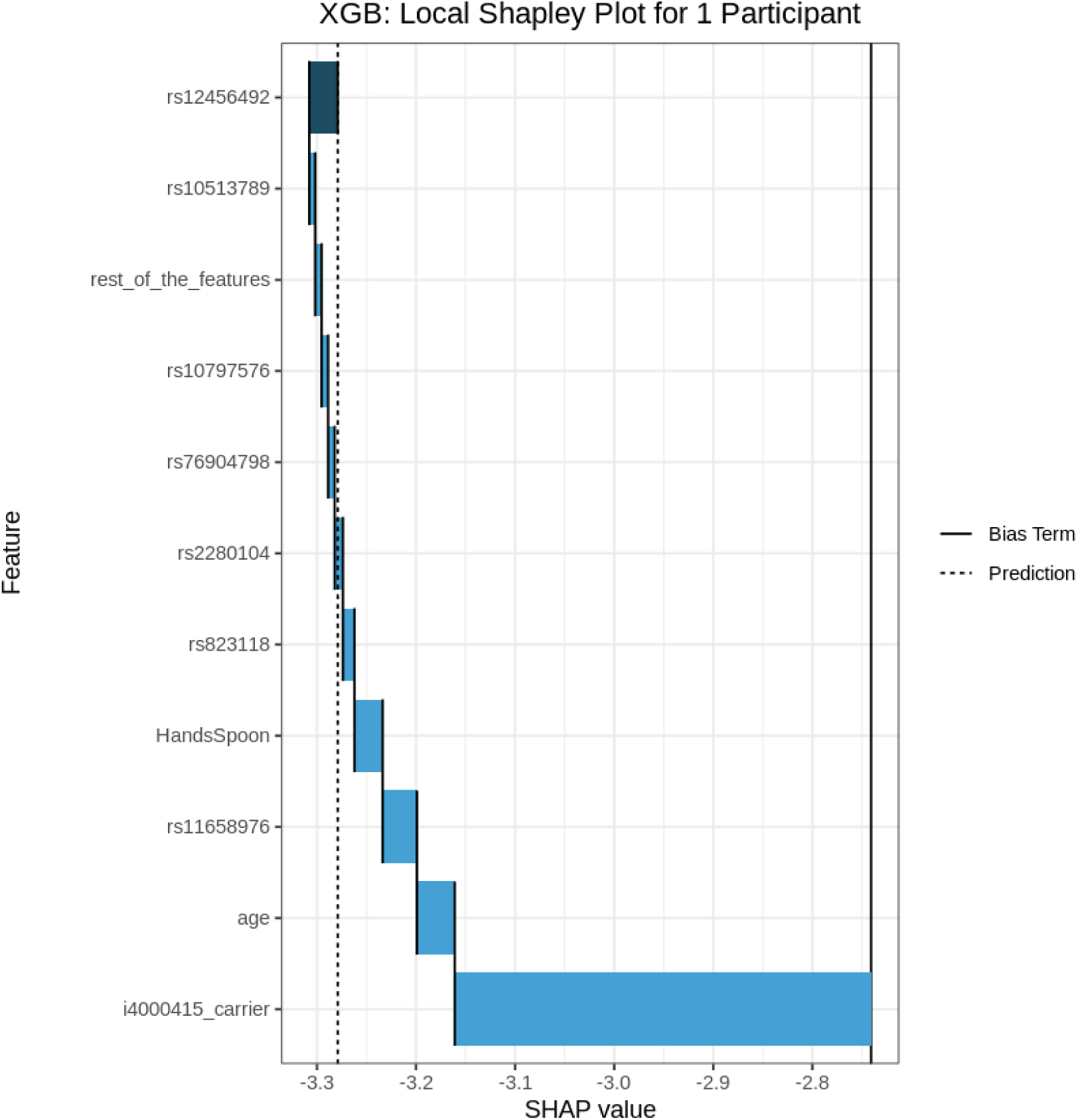

#### DNN

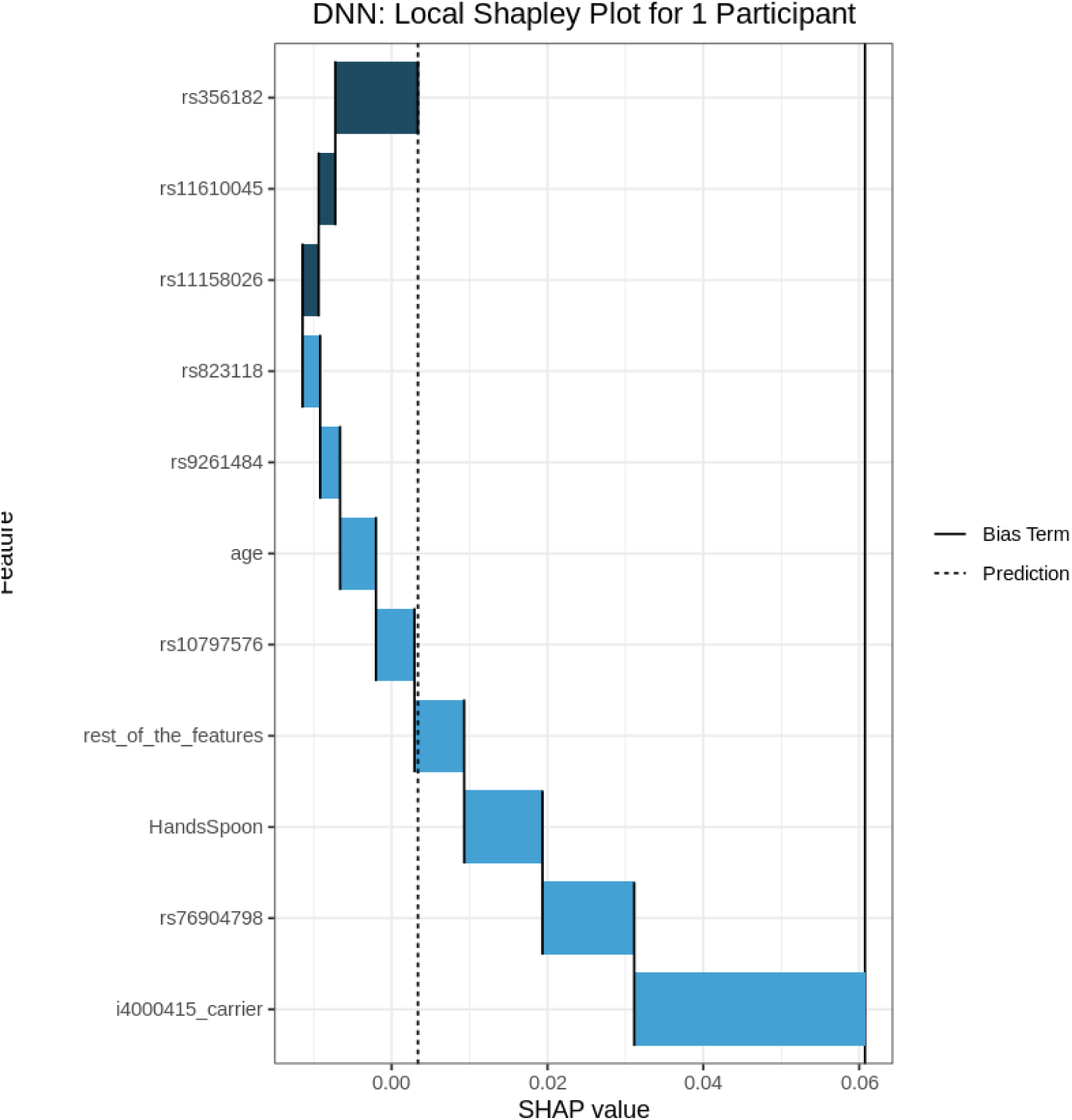

### Compare Predictions Across Models

#### Control

##### Build group 2

| Prediction by Model for 1 Control Participant | PD | Model |
| --- | --- | --- |
| 1 | 0 | deeplearning |
| 0.971 | 0.029 | ensemble |
| 0.967 | 0.033 | gbm |
| 0.967 | 0.033 | xgboost |
| 0.962 | 0.038 | gbm |
| 0.947 | 0.053 | gbm |
| 0.915 | 0.085 | gbm |

##### Build group 1

| Prediction by Model for 1 Control Participant | PD | Model |
| --- | --- | --- |
| 0.997 | 0.003 | deeplearning |
| 0.995 | 0.005 | ensemble |
| 0.978 | 0.022 | gbm |
| 0.975 | 0.025 | glm |
| 0.964 | 0.036 | drf |
| 0.964 | 0.036 | xgboost |
| 0.958 | 0.042 | xrt |

#### PD

##### Build Group 2

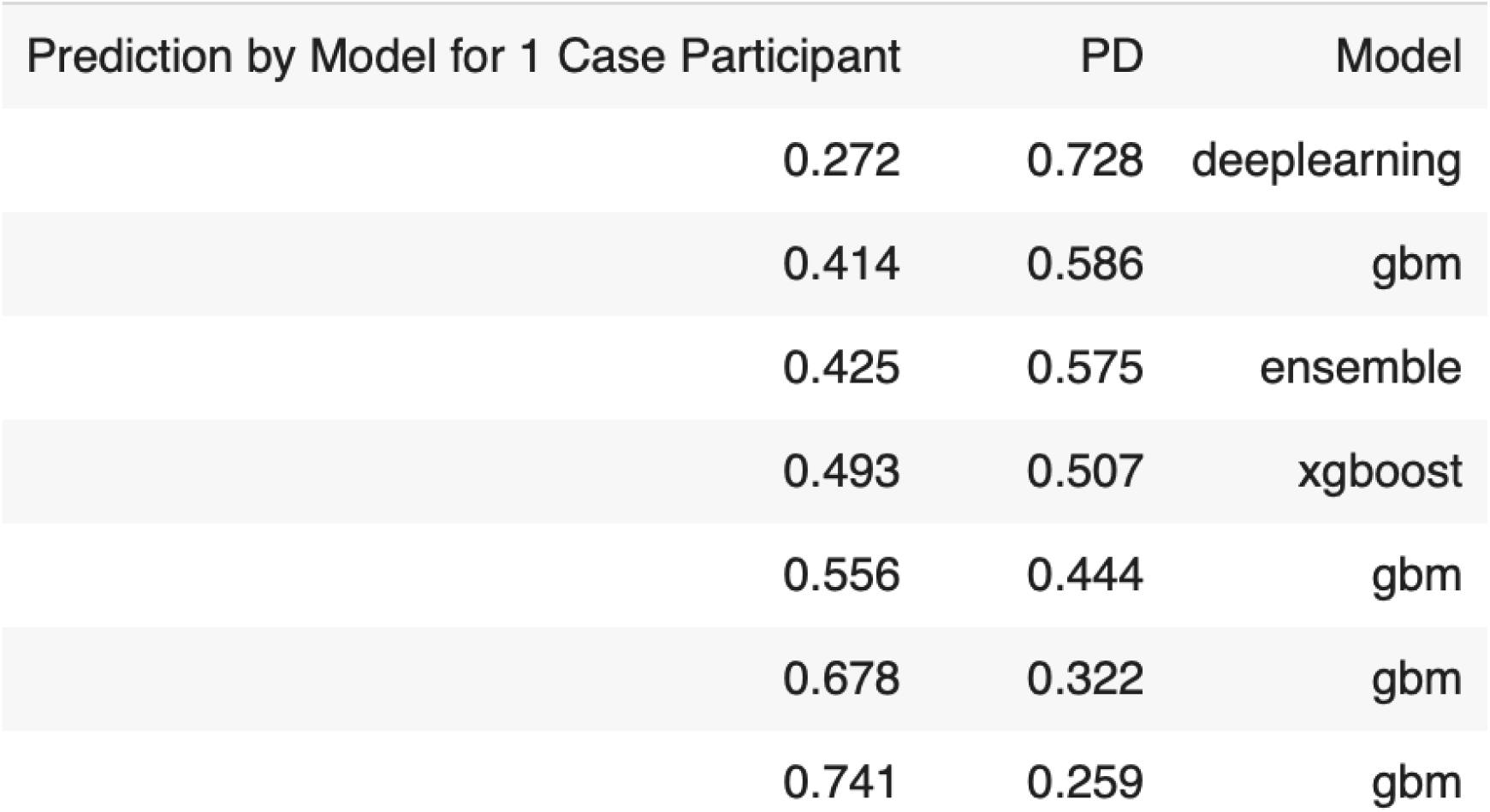

##### Build Group 1

| Prediction by Model for 1 Case Participant | PD | Model |
| --- | --- | --- |
| 0.077 | 0.923 | deeplearning |
| 0.381 | 0.619 | xgboost |
| 0.393 | 0.607 | glm |
| 0.452 | 0.548 | drf |
| 0.532 | 0.468 | ensemble |
| 0.625 | 0.375 | gbm |
| 0.722 | 0.278 | drf |

#### Generalized Low Rank Model (Feature Compression)

##### Evaluation

The GLRM-enhanced GBM model fit with learning rate annealing performed best, as shown below (on the unseen testing set):

| AUC | Gini | Log Loss | MCC |
| --- | --- | --- | --- |
| 0.902 | 0.805 | 0.116 | 0.727 |

Regarding the 280 control participants within the validation sample, the model incorrectly assigned a disease classification to 5 individuals, representing a 1.8% false-positive rate. Conversely, for the 17 participants with a clinical diagnosis of PD in this held-out set, the classifier failed to detect risk in 4 instances, resulting in a 23.5% false-negative rate.

### Variable Importance

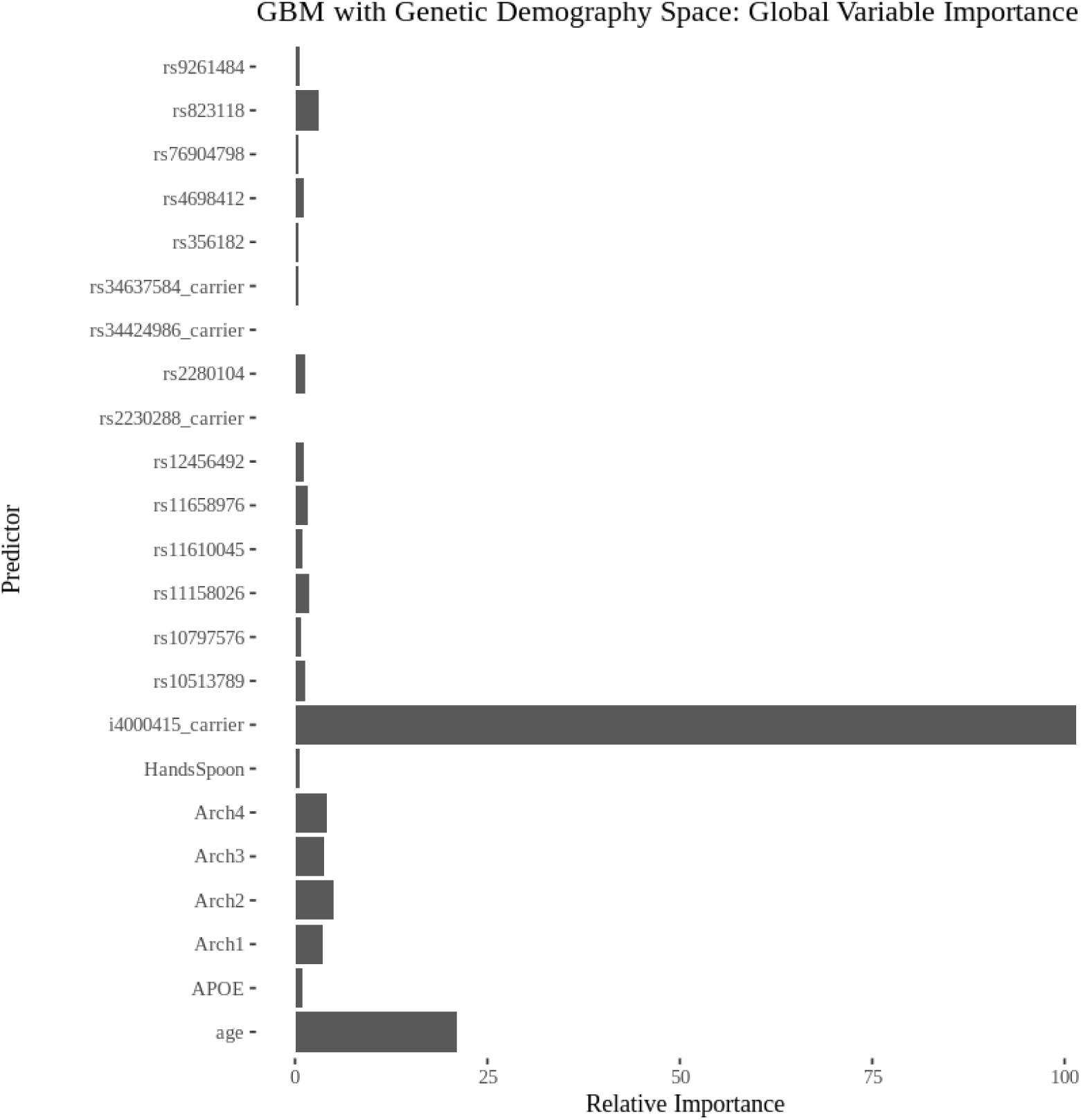

### Individual Conditional Expectations

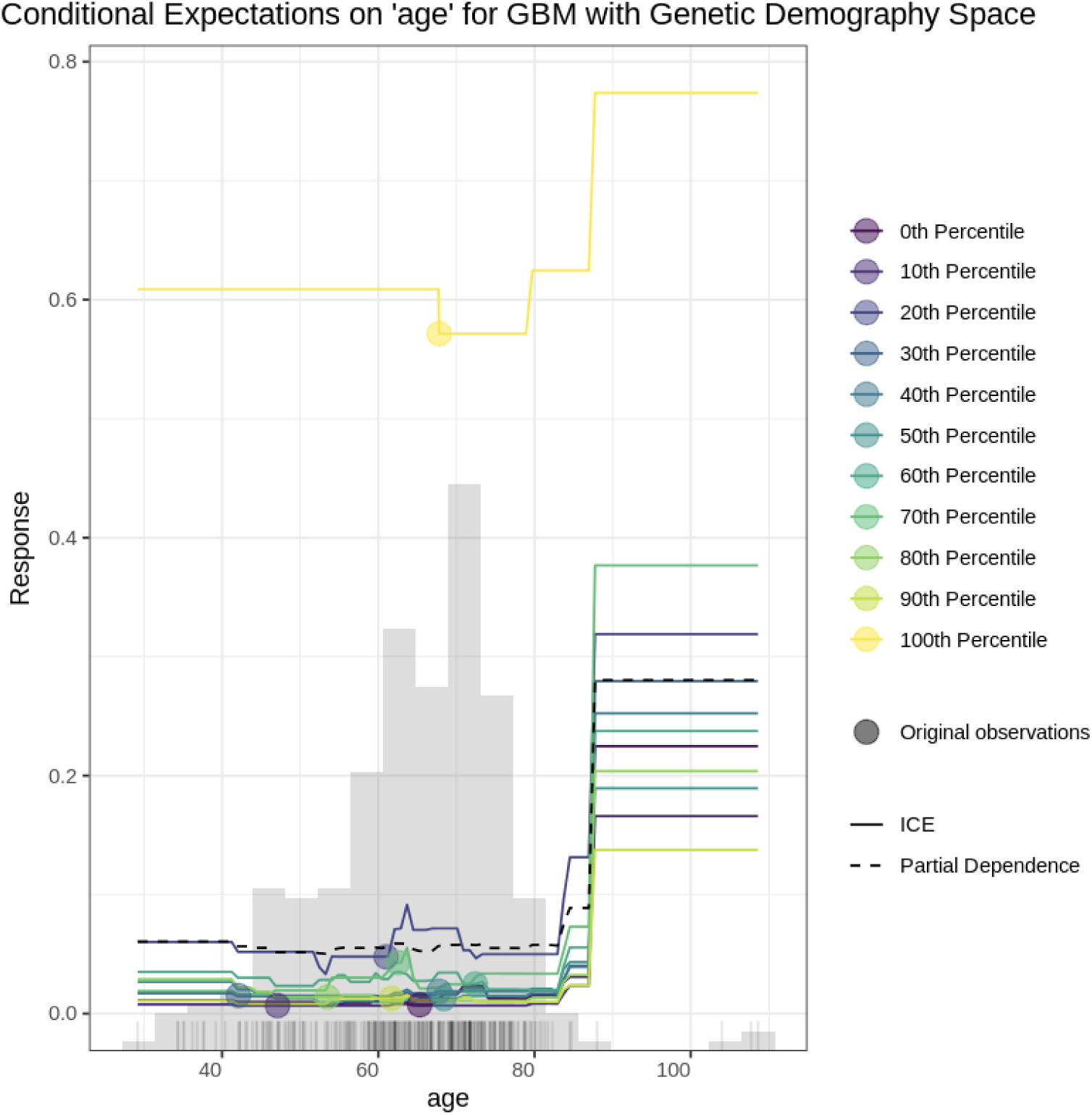

### Shap-Local

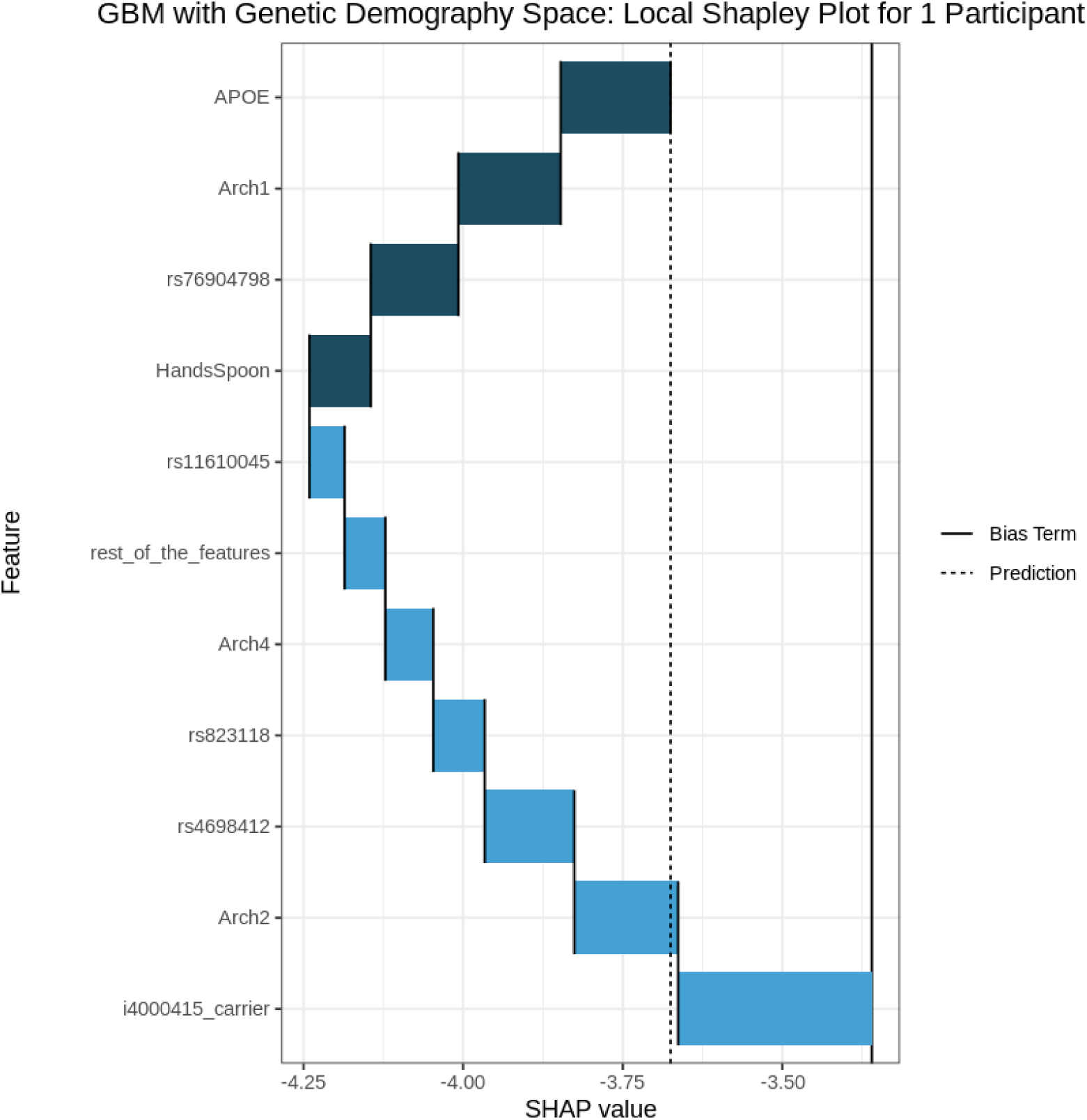

## Conclusion

While the traditional statistical model (Firth) provides *in-sample* predictive utility (AUC 0.905) and interval-bound point estimates, the ML approach provides ability to capture complex, non-linear interactions pushing the predictive power even higher (AUC 0.929) on *entirely unseen data*. In consideration of the estimated decreased risk factor for PD attributed to i4000415/N370S/GBA1 (risk variant carriers relative to non-carriers) in the Fox Insight analysis sample, in the interest of maintaining concern, conclude that this contradicts previous work suggesting risk variants of this genetic loci may be associated with *increased* PD risk in the population (Parlar, Grenn, Kim, Baluwendraat & Gan-Or, 2023). This provides reason to caution *inference* based on this finding in terms of population generalizability and instead to raise awareness of the intentional cohort enrichment - *while maintaining the utility of what is demonstrated as in-distribution predictive value*. The genetic targets employed as features for prediction are offered for this utility while their *a priori* inclusion is based on inferred biological basis in Parkinsononian phenomena.

Taken together, the classical statistical model and machine learning models represent a suite of deployable, *interpretable* classifiers trained on clinical diagnoses of PD that demonstrate the ability to discern sources of relevant, stable (within-person), predictive information. Machine learning model evaluation based on the Fox Insights participant sample *hold-out* partition (unseen during model training; ‘out-of-bag’) demonstrates *in-distribution generalizability*, however, it is unknown whether the suite of models are tenable among other participant groups (‘*out-of-distribution*’). The modeling process demonstrates replicable techniques that can be repurposed for other samples and even wider feature sets to support economical clinical assessment and long-horizon triage. This framework reveals a pivotal synthesis: though sophisticated ensemble methods effectively model intricate, non-linear dependencies, traditional statistical screening and rigorous baseline assignment are indispensable. Such grounding is required to ensure that algorithmic outputs do not misinterpret recruitment-driven structure, notably the GBA N370S enrichment, as biological signals. Finally, while other targeted genetic variants were deemed statistically non-zero in their *inferential* value with regard to Parkinsonian diagnosis in the participant sample, the moderate (relative to Bonferroni, for instance) FDR procedure protects us from inferential commitments based on the statistical model while allowing us to leverage biologically plausible genetic features for prediction.

## Discussion

While the GLRM compressed features did not improve the global predictive ability of the classifiers, an expanded GLRM fitting approach may yield different results. Additionally, this data-heterogeneity-capable, unsupervised modeling framework can become valuable with visualization of compressed dimensions in terms of the x-matrix (demography space) to identify clusters of participants from high-dimensional space as well as the y-matrix (feature space), where mixed data can be projected into low-dimensional archetypes, providing projections of canonical spread and clustering of continuous, numeric features along with every level of each factor variable included, for instance.

While the inclusion of a targeted genetic array provide specific sources of variance hypothesized to be associated with long-horizon Parkinsonian developments and yield valuable predictive insight, other data sources leveraging clinical imaging, additional biomarkers, and (participant) voice samples are relevant and may provide greater predictive ability and explanatory power from more subspaces of valuable variance. Twala (2025) provided multimodal PD classification models trained on *synthetic* participant groups, demonstrating how models trained on multiple kinds of data can bolster prediction and be applied to humans. Nevertheless, researchers and clinicians shouldn’t lose sight of the reductionist nature of the lenses built up from various data-driven modalities. The task of pushing our complementary tools as far as they can go is before us, but so is keeping at the forefront - that the humans we wish to serve are invariably more nuanced in their experiences and trajectories than our instruments can imagine.

## Data Availability

All data produced are available online at: https://foxden.michaeljfox.org

## Appendix A.

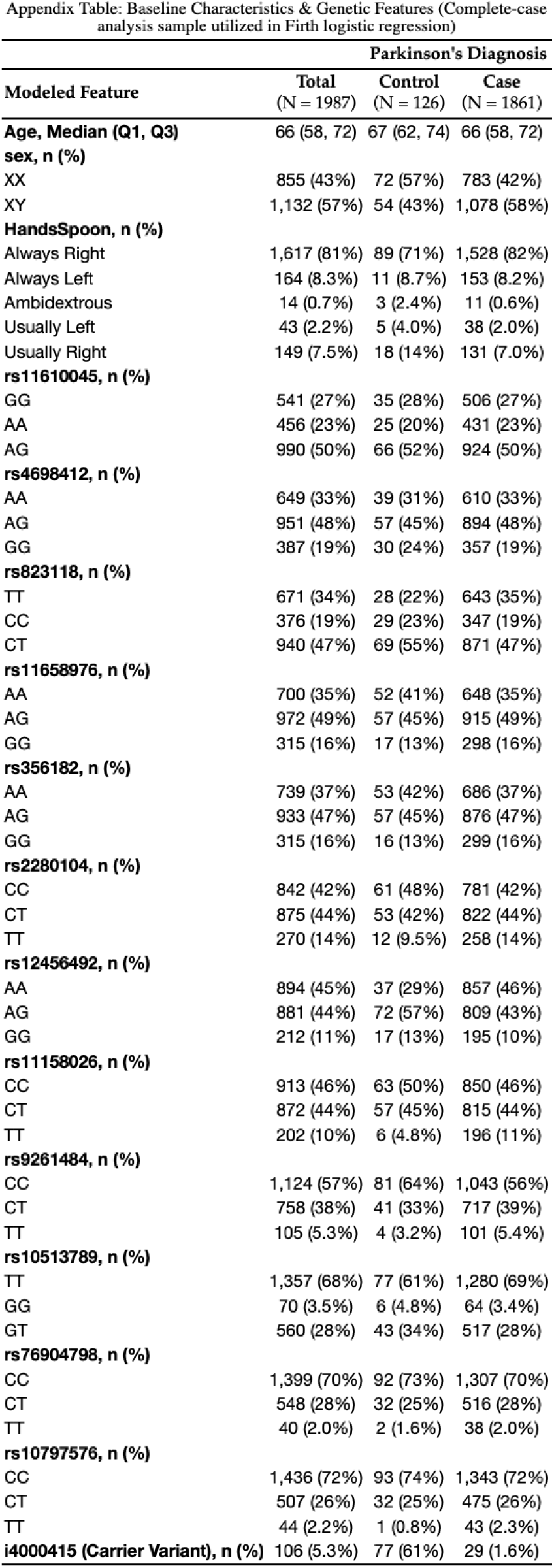

## Footnotes

1 Given the nature of the participant sample, the objective, here, should be maintained that causal genetic variant effects *may not be estimated for the population* while predictive insights pertinent to the defined Fox Insights sample are provided (sample-specific associations rather than estimates of biological magnitude).

## Notes

### Competing Interest Statement

The authors have declared no competing interest.

### Author Declarations

All data are individual-level, de-identified.

